# The overlooked impact of background diet and adherence in nutrition trials

**DOI:** 10.1101/2025.02.26.25322933

**Authors:** Javier I. Ottaviani, Hagen Schroeter, Dennis M. Bier, John W. Erdman, Howard D. Sesso, JoAnn E. Manson, Gunter G. C. Kuhnle

## Abstract

Randomized controlled trials in nutrition (RCTN) face unique challenges, including the considerable influence of the background diet and the challenge of assuring intervention adherence by participants. The impact of these factors on the outcome of RCTNs has been difficult to quantify, but nutritional biomarkers represent a valuable tool to address these challenges. Using flavanols as a model dietary intervention and a set of recently validated flavanol biomarkers, we here investigated the impact of background diet and adherence on the outcomes of the COcoa Supplement and Multivitamin Outcomes Study (COSMOS, NCT 02422745). We found that 20% of participants in the placebo and cocoa-extract intervention arms had a flavanol background intake as high as the intervention, and only 5% did not consume any flavanols. Approximately 33% of participants in the intervention group did not achieve expected biomarker levels from the assigned intervention – more than the 15% estimated with pill-taking questionnaires usually implemented in RCTN. Taking these factors into account resulted in a larger effect size for all observed endpoints (HR (95% CI)) estimated using intention-to-treat vs. per-protocol vs. biomarker-based analyses: total cardiovascular disease (CVD) events 0.83 (0.65; 1.07); 0.79 (0.59; 1.05); 0.65 (0.47; 0.89) – CVD mortality 0.53 (0.29; 0.96); 0.51 (0.23; 1.14); 0.44 (0.20; 0.97) – all-cause mortality 0.81 (0.61; 1.08); 0.69 (0.45; 1.05); 0.54 (0.37; 0.80) –– major CVD events 0.75 (0.55; 1.02); 0.62 (0.43; 0.91); 0.48 (0.31; 0.74). These results highlight the importance of taking background diet and adherence into consideration in RCTN to obtain more reliable estimates of outcomes through nutritional biomarker-based analyses.

## Introduction

Randomized controlled trials (RCTs) are generally considered the gold standard for the scientific study of drugs, treatments and dietary interventions in medical ^1-4^ and nutrition research ^5-7^, respectively. However, nutrition trials (RCTN) face unique challenges ^8-10^ that are often overlooked. In particular, the impact of background diet of participants on trial endpoints and the objective assessment of adherence are key to the interpretation of trial outcomes. Unlike in medical RCTs where the uncontrolled and unknown exposure to a test drug or treatment can almost never occur, participants in RCTN will almost always be exposed to foods, nutrients, or dietary constituents that are similar to or indistinguishable from the study intervention ^8^. Very often this exposure from background diet cannot be quantified by investigators and it is therefore not included in outcomes analyses ^11^. The same applies to adherence, which can be more easily monitored with pharmacological interventions where specific and unambiguous biomarkers exist ^12-14^, whereas most RCTN have to rely exclusively on self-reported information ^15-20^ – approaches that carry a higher risk of misclassification of adherence. Taken together, these two challenges can significantly affect the outcomes and mask differences between intervention and control groups, leading to incorrect interpretations. Considering the cost, efforts, and high impact of, especially large-scale RCTN, it is important to quantify the potential error introduced by these two often overlooked factors, and to identify and use better methods to address these limitations. This will allow to generate RCTN outcomes of higher scientific rigor and evidence quality, more comparable to RCT in medical research, and contribute to increase reliability and confidence in nutrition research by providing clearer messages to practitioners and the general public

Validated nutritional biomarkers provide the opportunity to address both challenges in RCTN ^21,22^. Nutritional biomarkers measure the systemic presence of the dietary compound under investigation and thereby can provide objective information on both, background diet and adherence to the intervention. However, because there are so few nutritional biomarkers that have been validated ^22^, integration in large-scale RCTN has been rare and the actual impact is currently unknown.

Flavanols are a group of food bioactives for which a beneficial effect on health has been established^23,24^. The particular group of flavanols present in cocoa, namely cocoa flavanols (CF), have been investigated recently in a large RCTN, COSMOS (COcoa Supplement and Multivitamin Outcomes Study), which showed that CF intake reduces cardiovascular disease (CVD) risk ^25^. There exist two validated biomarkers for the intake of flavanols, namely urinary 5-(3□,4□-dihydroxyphenyl)-γ-valerolactone metabolites (gVLM_B_) and structurally related (–)-epicatechin metabolites (SREM_B_) ^26,27^. These biomarkers have been successfully applied in observational studies to assess flavanol intake ^28^, including the intake of those specific flavanols that are part of the CF evaluated as part of COSMOS intervention ^26,27^.The COSMOS trial included the collection of spot urine samples at baseline and during follow-up allowing objective, biomarker-based assessments of both participants’ background diets and adherence to the intervention using gVLM_B_ and SREM_B_ as validated nutritional biomarkers ^26,27^. Hence, CF and COSMOS represent an excellent model to assess the impact that dietary background and adherence could have on the outcomes of a large RCTN.

## Materials and Methods

### Study design

This study consisted of a post-hoc, secondary analysis of a subcohort (n=6,532) of COSMOS (NCT02422745; Figure S1 in Supplementary Information), a recently completed RCTN in 21,442 participants (including 8776 males ≥60 y and 12,666 females ≥65 y) in the US ^25^. The interventions included capsules containing cocoa extract that provided 500 mg/d cocoa flavanol (CF) or placebo, and a Centrum Silver daily multivitamin or placebo (provided by Pfizer Consumer Healthcare, now Haleon) in a 2×2 factorial design (see Supplementary Materials for details). Participants of the COSMOS Biomarker Cohort provided spot urine samples at baseline during the run-in phase of the study prior to randomization and again at 1, 2 and/or 3-year follow-up. Participants completed a self-reported adherence assessment every 6 months by answering a series of questions related to the number of days taking study pills ^29^. Enrolment period for the entire group of participants in COSMOS extended from April 2016 to March 2018, and the intervention was completed on December 31, 2020, with a median treatment period of 3.6 years. All participants provided written informed consent, and study approvals were obtained by the Institutional Review Board (IRB) at Mass General Brigham.

### Flavanol biomarker quantification

Urinary levels of 5-(3□,4□-dihydroxyphenyl)-γ-valerolactone metabolites (gVLM_B_) and structurally related (–)-epicatechin metabolites (SREM_B_) were used as biomarkers of flavanol intake. gVLM_B_ represented the sum of the urinary concentrations of 5-(4□-hydroxyphenyl)-γ-valerolactone-3□-sulfate and 5-(4□-hydroxyphenyl)-γ-valerolactone-3□-glucuronide (5-(3□-hydroxyphenyl)-γ-valerolactone-4□-sulfate and 4□-glucuronide were not included as these represented minor metabolites ^30^ and did not affect the performance of gVLM_B_ as a biomarker ^28^). SREM_B_ represented the sum of the urinary concentrations of (–)-epicatechin-3□-glucuronide, (–)-epicatechin-3□-sulfate and 3□-O-methyl(–)-epicatechin-5-sulfate. gVLM_B_ and SREM_B_ were validated as nutritional biomarkers of flavanol intake ^26,27^, and were successfully implemented in observational studies ^28^. While gVLM_B_ informs on the intake of flavanols in general (particularly flavanols including a catechin or epicatechin moiety in their structure), SREM_B_ is a specific biomarker of the intake of (–)-epicatechin, one of the main bioactive flavanol compounds in the 500 mg/d CF tested in COSMOS. SREM_B_ and gVLM_B_ have different systemic half-lives ^30^, thus a combination of both biomarkers allows capturing different periods after flavanol intake. gVLM_B_ and SREM_B_ were quantified using validated LC-MS methods ^28^ and performance of the quantification are provided in Supplementary Information. Unadjusted biomarker concentrations were used as urinary creatinine is associated with CVD risk ^31^ and specific gravity did not materially change the outcome (see Supplementary Data for further details).

### Flavanol intake assessment using flavanol biomarkers

The quantification of gVLM_B_ and SREM_B_ in spot urine serve as concentration biomarkers of flavanol intake and thus, these biomarkers could not be used to determine absolute flavanol intake ^21^. Instead, we have used gVLM_B_ and SREM_B_ concentrations to classify participants into having flavanol intakes either below, or equal and above the 500 mg/d tested in COSMOS. Thresholds for gVLM_B_ and SREM_B_ concentrations were derived from a dose-escalation study conducted as part of the validation of these flavanol biomarkers ^26,27^, in which gVLM_B_ and SREM_B_ were expressed in µM concentrations in urine. Considering the inter-individual variability in biomarker levels and to reduce the likelihood of false negatives, respective thresholds were conservatively defined as the bottom 95% CI limit of the expected concentration of gVLM_B_ and SREM_B_ after the intake of 500 mg of flavanols (**Figure 1**), using a linear regression model with log_2_-transformed concentration data. These thresholds were thus determined as 18.2 µM for gVLM_B_ and 7.8 µM for SREM_B_. Given the importance of thresholds selected on the outcomes of this study, we conducted sensitivity analyses using different thresholds for gVLM_B_ and SREM_B_ and investigate their impact on estimated outcomes.

**Figure 1.**
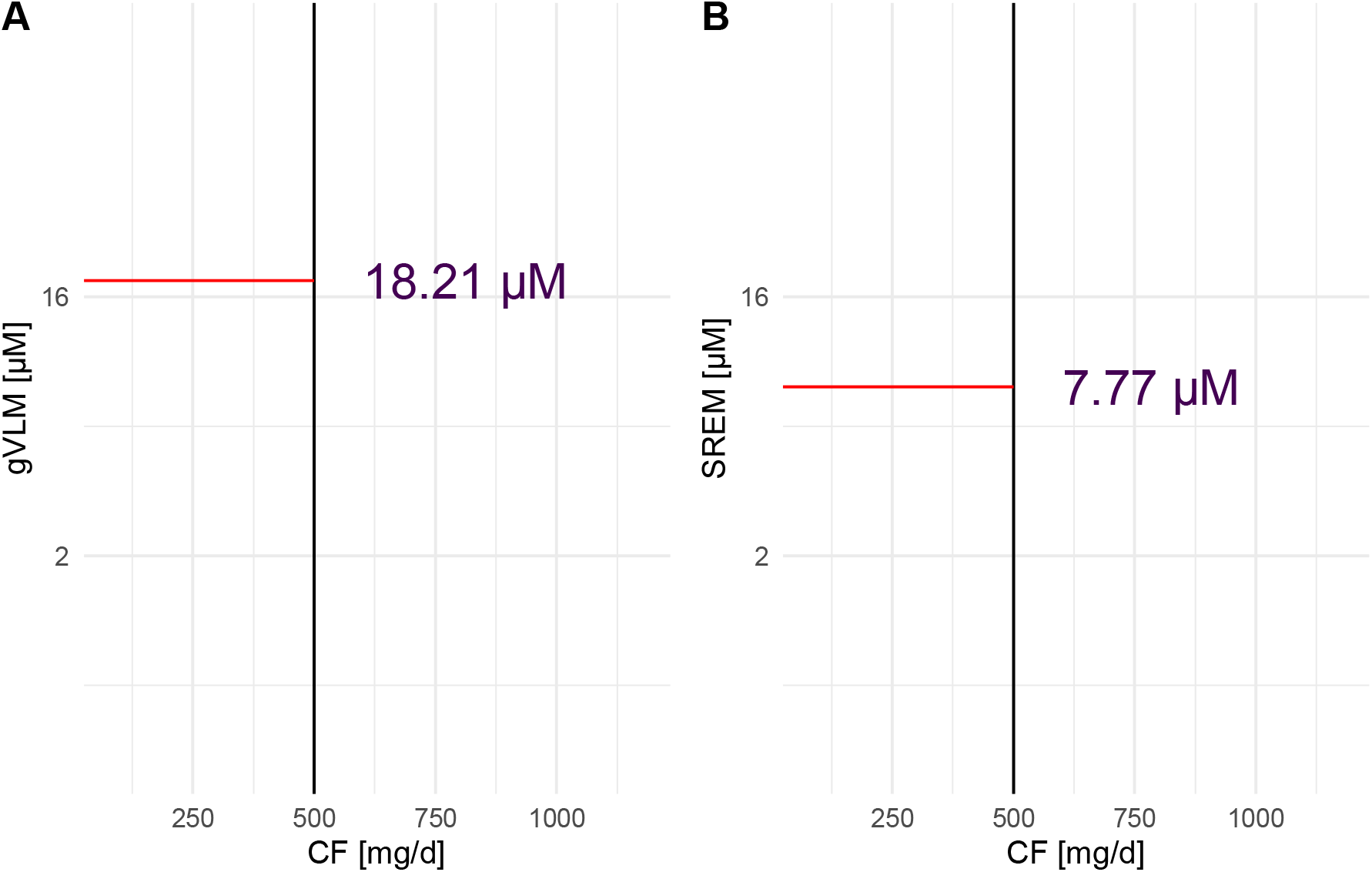
Concentration of gVLM_B_ (A) and SREM_B_ (B) in urine as a function of the amount of cocoa flavanols (CF) consumed. Flavanol biomarker data were log-transformed. gVLM_B_ and SREM_B_ thresholds were defined as the bottom 95%CI limit of the expected concentration of gVLM_B_ and SREM_B_ after the intake of 500 mg of CF. Data were obtained from data collected previously ^26, 27^. gVLM_B_: 5-(3□,4□-dihydroxyphenyl)-γ-valerolactone metabolites; SREM_B_: structurally related (–)-epicatechin metabolites.

### Endpoints

Endpoints of this study were i) total cardiovascular disease (CVD) events, the primary outcome in the COSMOS main trial that included myocardial infarction (MI), stroke, coronary revascularization, cardiovascular mortality, carotid artery surgery, peripheral artery surgery, and unstable angina requiring hospitalization; ii) CVD mortality, a secondary outcome in the COSMOS main trial; ii) all-cause mortality, a secondary outcome in the COSMOS main trial; and iv) major CVD events (MI, stroke, and CVD mortality), which are a recognized CVD outcomes, but was not included among pre-registered COSMOS outcomes. Participants reporting an outcome signed a release form to request related medical records for evaluation and processing according to standardized WHI and BWH study procedures. Self-reported primary and secondary outcomes were confirmed by medical record review by a committee of physicians and investigators blinded to treatment assignment. Further details on endpoint adjudication were published previously ^25^. Median follow-up time was 3.6 years.

### Statistical analysis

All analyses were conducted with R version 4.3.2 ^32^ in RStudio 2023.12.1 using the packages *rms* ^33^ for regression analyses and *ggplot2* ^34^ for the generation of graphics. Missing values were assumed to be missing at random and imputed using multiple imputation (including Nelson-Aalen estimator ^35^). Unless indicated otherwise, results are shown with 95% confidence intervals (CI). Associations between endpoints and study groups were investigated using Cox proportional hazards models. The proportional hazard assumption was tested and confirmed with the *cox*.*zph* function. Statistical models were selected *a priori* based on analyses conducted previously ^25^. Continuous variables were transformed using restricted cubic splines with 3 knots (outer quantiles 0.1 and 0.9). Model 1 was adjusted by sex, age at randomization and recruitment cohort (Women’s Health Initiative or other). Model 2 was further adjusted by BMI (kg/m^2^), smoking status (never, ever, current), and aspirin use (yes/no). Model 3 was additionally adjusted by alternative Healthy Eating Index (aHEI, continuous), family history of hypertension, high cholesterol and stroke (categorical) and history of hypertension (categorical). Model 4 was like model 2, but additionally adjusted by randomization (Placebo or CF intervention), model 5 was like model 2, but additionally adjusted by aHEI. These different models were used to conduct sensitivity analysis of the outcomes obtained. All results were shown as estimated hazard ratio (HR) (95% CI) between control and intervention. For per protocol analyses, participants were censored on the date when participants self-reported missing either more than 8 pills per month or that they were unsure how many pills they have missed, or began outside non-study use of cocoa supplement. Cumulative incidence plots were created using the *survplot* function and data from model 2 (including age at randomization, sex, BMI, smoking status, aspirin use and recruitment cohort).

### Data and code availability

Code is available from https://gitlab.act.reading.ac.uk/xb901875/biomarker-based-intervention. Data and associated documentation will be available to users only under a data-sharing agreement. Details on the availability of the study data to other investigators will be on our study website at https://cosmostrial.org/.

## Results

### COSMOS Biomarker cohort

A total of 6,509 out of the 21,442 participants randomized in COSMOS provided spot urine samples for the quantification of flavanol biomarkers (gVLM_B_ and SREM_B_; see Figure S1 and Table S1 for more details).

### Background diet and adherence assessment using flavanol biomarkers

Biomarker concentrations at baseline and follow up in the COSMOS Biomarker cohort are shown in Figure 2. At baseline, biomarker concentration in approximately 20% of participants were consistent with flavanol intake of at least 500 mg/d as part of their background diet, with little differences between participants on the intervention and control group (Table 1). Only 5% of participants had biomarker concentrations below the limit of quantification of the analytical method, and thus indicate a negligible or very low exposure to flavanols from the background diet.

**Table 1.** Number of participants with flavanol biomarker levels consistent with a flavanol intake of 500 mg/d or higher at baseline and follow-up.

| | Biomarker-estimated flavanol intake ≥ 500 mg/d | |
| --- | --- | --- |
|  | Baseline (background diet) | Follow-up (adherence) |
| Participants with spot urine at baseline (n=6,509) | 1,249 (19%) | — |
| Placebo (n=3,257) | 603 (19%) | — |
| CF intervention (n=3,252) | 646 (20%) | — |
| Participants with spot urine at baseline and follow-up (n=2,051) | 414 (20%) | 881 (43%) |
| Placebo (n=991) | 186 (19%) | 175 (18%) |
| CF intervention (n=1,060) | 228 (22%) | 706 (67%) |

**Figure 2.**
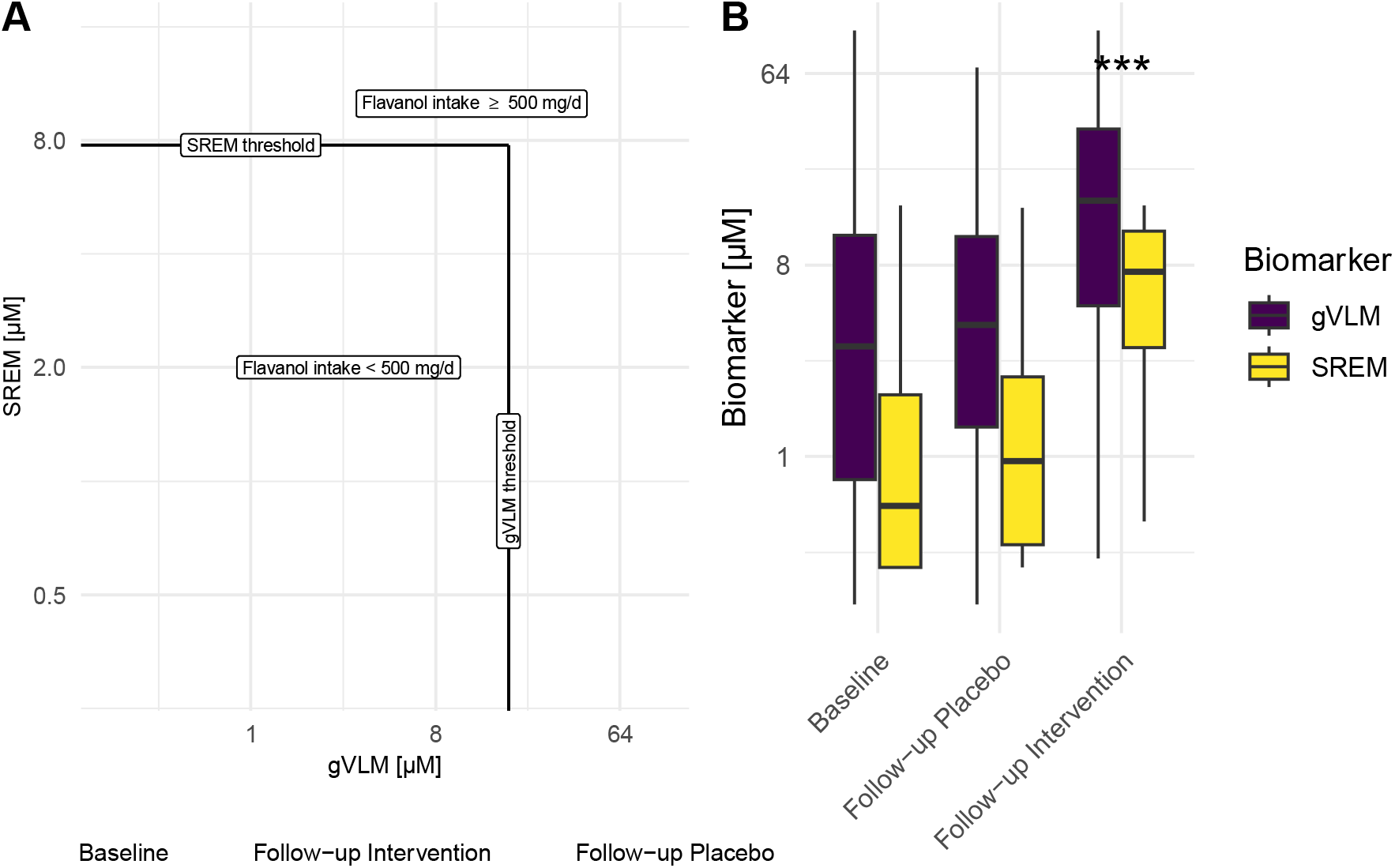
**A)** Concentration of SREM_B_ and gVLM_B_ in individual participants (n=6,509) at baseline and follow-up in intervention and placebo groups. Black line represents thresholds for SREM_B_ and gVLMB. Participants having gVLM_B_ or SREM_B_ levels above the corresponding thresholds were considered to have a flavanol intake equal or higher than 500 mg/d. B) Concentration of SREM_B_ and gVLM_B_ in participants (n=6,509) at baseline and follow-up in intervention and placebo groups. Data are shown as box-plots, with the box representing the inter-quartile range. Biomarker concentration at follow-up in the intervention group were significantly higher (Tukey’s Honestly Significant Difference).

Among 2,051 participants with follow-up spot urine samples, 67% of those assigned to the intervention group had a flavanol consumption consistent with the intake of at least 500 mg/d of flavanols (Table 1) and were thus considered adherent to study pill intake. This figure was lower than the adherence determined using self-reported methods, which was 85% ^25^.

### Biomarker-based intervention groups

We used flavanol biomarker data to identify participants as either part of the *biomarker active* or *biomarker control* groups, analogous to the intervention-based *intervention* and *control* group. *Biomarker active* participants had a biomarker estimated flavanol intake of at least 500 mg/d either at baseline or follow-up. Those in the *biomarker control* group did not achieve these biomarker concentrations at baseline and at follow up. Participants without a follow up sample in the CF intervention group who had a background diet providing less than 500 mg/d of flavanols were excluded from the analysis (n=1,774; 27% of Biomarker Cohort) as adherence could not be determined in this group.

Only 62% of participants in the *biomarker active* group had been randomized into the intervention group, whereas 10% of participants in the *biomarker control* group had been randomized into the intervention. In addition to differences in the number of participants receiving placebo and intervention in the biomarker active and biomarker control groups, additional differences in the baseline characteristics of these participants are shown in Table S2.

### CVD events in biomarker-based intervention groups

Table 2 shows the associations between intervention groups and disease risk among those in the COSMOS Biomarker cohort. There were substantial differences between HRs when using biomarker-based groups and the randomized groups. The *biomarker active* group showed statistically significant reduced risks for all endpoints, while the associations were weaker when using groups based on randomization assignment. In the intention-to-treat (ITT) analysis, which compares intervention and placebo and does not take adherence and background diet into consideration, a significant reduction in risk was found only for CVD mortality, consistent with the main COSMOS trial findings ^25^. Taking self-reported adherence into consideration (per protocol analysis, PP), we also found significant risk reductions for major CVD events and all-cause mortality, but not total CVD events.

**Table 2.** HRs and 95% CIs for cardiovascular (CVD) outcomes, according to randomized assignment in intention-to-treat (ITT) and per-protocol (PP) analyses and according to biomarker-based groups. Summary statistics were from Cox regression models adjusted by age, sex, BMI, smoking status, recruitment cohort and aspirin use (model 2). CIs were not adjusted for multiple comparisons.

|  | Intervention-based groups |  | Biomarker-based groups |
| --- | --- | --- | --- |
|  | ITT<sup>†</sup> (n=6,532) | PP<sup>‡</sup> (n=6,532) | Biomarker (n=4,735) |
| All-cause mortality | 0.81 (0.61; 1.08) | 0.69 (0.45; 1.05) | 0.54 (0.37; 0.80) |
| Total CVD events | 0.83 (0.65; 1.07) | 0.79 (0.59; 1.05) | 0.65 (0.47; 0.89) |
| CVD mortality | 0.53 (0.29; 0.96) | 0.51 (0.23; 1.14) | 0.44 (0.20; 0.97) |
| Major CVD events | 0.75 (0.55; 1.02) | 0.62 (0.43; 0.91) | 0.48 (0.31; 0.74) |

Total CVD events is a composite of myocardial infarction, stroke, CVD death, coronary artery bypass graft and percutaneous coronary intervention, unstable angina including hospitalization, carotid artery surgery, and peripheral artery surgery. Major CVD events was a composite of myocardial infarction, stroke, and CVD death. CVD, cardiovascular disease; <sup>†</sup>Intention to treat; <sup>‡</sup>per-protocol

The cumulative incidence curves for total CVD events in the COSMOS trial and in the COSMOS biomarker cohort when using groups based on randomization showed a divergence in hazard rates from approximately one year of follow-up (Figure 3B). In contrast, when using *biomarker-based* intervention groups, divergence starts earlier (Figure 3A). This difference could be the consequence of baseline dietary flavanols mediating an effect in those participants that already had biomarker levels consistent with an intake of flavanol equal or above 500 mg/d before commencement of COSMOS trial.

**Figure 3.**
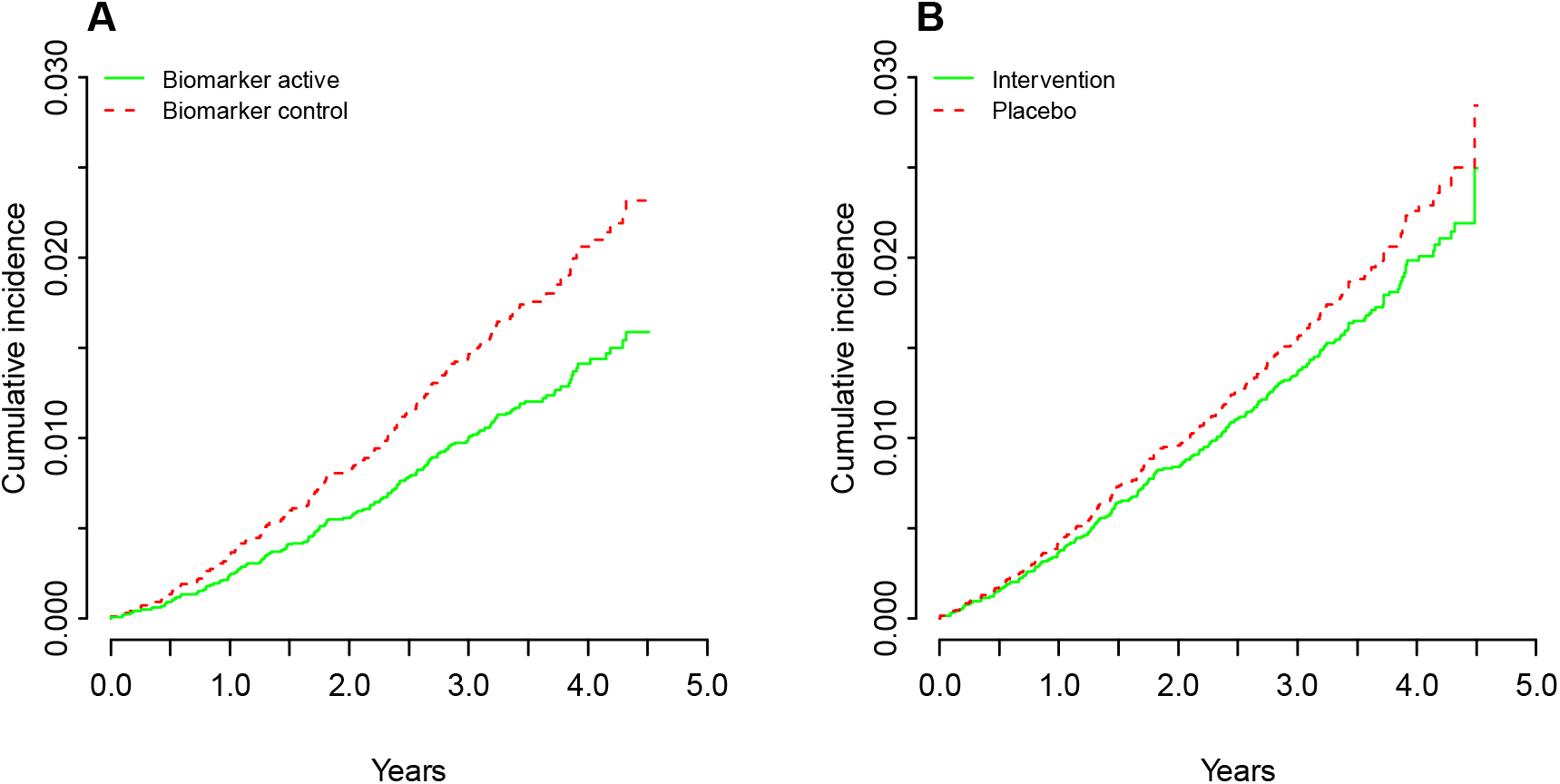
Cumulative incidence of total cardiovascular disease (CVD) events according to biomarker-based groups (A) and randomized assignment in per-protocol analysis (B). Results shown are adjusted for age at randomization (75), sex (male), BMI (25 kg/m^2^), smoking status (never), aspirin use (no) and recruitment cohort (WHI). Total CVD events was a composite of myocardial infarction, stroke, CVD death, coronary artery bypass graft and percutaneous coronary intervention, unstable angina including hospitalization, carotid artery surgery, and peripheral artery surgery.

### Sensitivity analyses

A series of sensitivity analyses were conducted to determine robustness of the findings for biomarker-based intervention groups (Tables 3). These analyses showed that adjusting for diet quality did not alter the results obtained, suggesting that flavanol intake, rather than a broader beneficial dietary pattern, may be responsible for the differences in CVD events between biomarker-based groups. Additional adjustment by randomization group attenuated the estimated association for major cardiovascular events), which is not surprising as most participants in the *biomarker active* group are also in the CF intervention group.

**Table 3.** Sensitivity analyses. HRs and 95% CIs for cardiovascular disease (CVD) outcomes using biomarker-based groups and different models. Summary statistics from Cox regression models show HRs comparing *biomarker control* and *biomarker active* group for each outcome. CIs were not adjusted for multiple comparisons.

|  | Model 1† | Model 2† | Model 3† | Model 4† | Model 5† |
| --- | --- | --- | --- | --- | --- |
| All-cause mortality | 0.52 (0.35; 0.77) | 0.54 (0.37; 0.80) | 0.57 (0.39; 0.85) | 0.65 (0.41; 1.02) | 0.55 (0.37; 0.82) |
| Total CVD events | 0.64 (0.47; 0.88) | 0.65 (0.47; 0.89) | 0.68 (0.49; 0.93) | 0.70 (0.48; 1.02) | 0.66 (0.48; 0.90) |
| CVD mortality | 0.42 (0.19; 0.93) | 0.44 (0.20; 0.97) | 0.46 (0.21; 1.02) | 0.64 (0.26; 1.54) | 0.45 (0.20; 0.98) |
| Major CVD events | 0.48 (0.31; 0.73) | 0.48 (0.31; 0.74) | 0.50 (0.33; 0.78) | 0.55 (0.33; 0.91) | 0.49 (0.32; 0.75) |

Given the importance of thresholds selected to define the biomarker-based CF active and control groups, we also investigated the impact of using different thresholds for gVLM_B_ and SREM_B_ to identify participant with flavanol intake of at least 500 mg/d. We have estimated associations between biomarker status (above threshold for gVLM_B_ or SREM_B_ [biomarker active] vs. below threshold for gVLM_B_ and SREM_B_ [biomarker control]) for thresholds varying between 1 µM and 50 µM for gVLM_B_ and 1 µM and 30 µM for SREM_B_. The results show that thresholds for gVLM_B_ varying between 3 to 20 µM and thresholds for SREM_B_ varying between 5 to 30 µM would not have resulted in a material change of observed estimates (Figure 4). Thus, the thresholds of 18.2 µM for gVLM_B_ and 7.8 µM for SREM determined for this study fall well within these ranges, and even considering potential inaccuracies in the approach chosen to determine the thresholds used in this study would have not changed the outcomes of this analysis.

**Figure 4.**
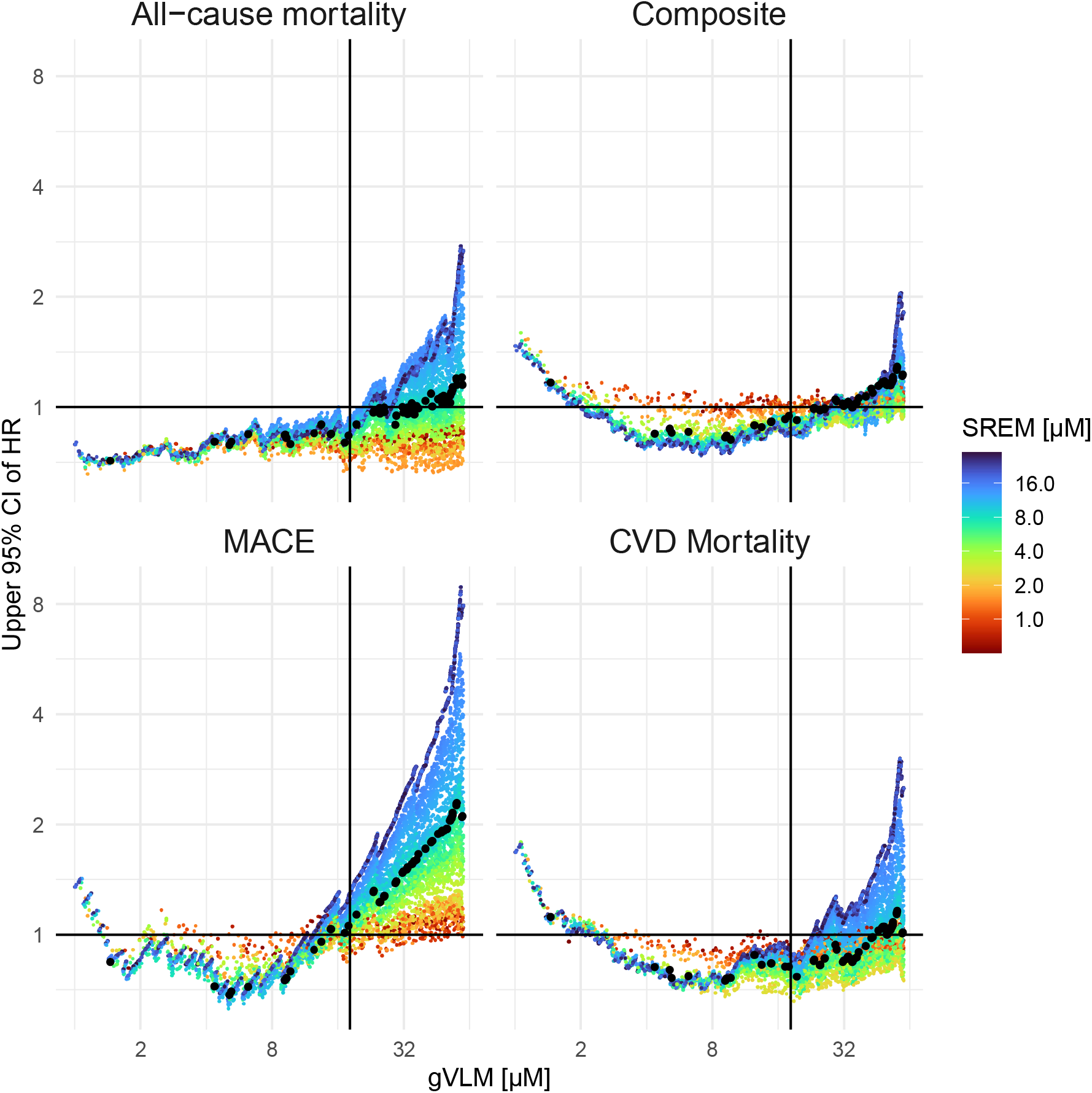
Comparison of 95% upper confidence limit of HR for different CVD events between biomarker-active and biomarker-control group when using different concentration thresholds. Summary statistics were from Cox regression models by age, sex, BMI, smoking status, and aspirin use. CIs were not adjusted for multiple comparisons. CVD events included total CVD events (which was a composite of myocardial infarction, stroke, CVD death, coronary artery bypass graft and percutaneous coronary intervention, unstable angina including hospitalization, carotid artery surgery, and peripheral artery surgery), CVD deaths, all-cause deaths and major adverse CVD events (which was a composite of myocardial infarction, stroke, and CVD death). Vertical lines and black dots indicate the gVLM and SREM thresholds used in this study. Horizontal line shows HR of 1.

## Discussion

In this study, we investigated two main limitations of RCTs in nutrition: background diet and adherence, and how these limitations could affect the outcomes interpretation of such studies. For this, we used nutritional biomarkers to objectively assess background diet and adherence, and using flavanols, flavanols biomarkers and the COSMOS trial ^25^ as a model system ^24, 31^. Our results show that both background diet and adherence have a substantial impact on the results of RCTNs:

1. One in five participants in both the placebo and intervention arms were already consuming flavanols as part of their background diet in amounts that were similar or higher to those tested in the intervention arm, and only one in twenty participants were not consuming any flavanols at baseline.
2. Approximately one third of participants assigned to the intervention did not attain the flavanol levels as set forth in this study for this intervention.
3. Considering both biomarker-estimated dietary background and adherence to study intervention resulted in insights on the effect size of the intervention that are more objective, reliable, and less impacted by methodological limitations compared to the general approach in RCTNs, which neither consideres background intake nor biomarker-estimated adherence.

The use of biomarkers allows the objective assessment of intake as part of the background diet and adherence to the intervention. This approach offers significant benefits compared to methods based on dietary and adherence questionnaires and food content databases, which are not only subjective in nature but known to incur significant levels of uncertainty ^36,37^. In contrast to biomarkers that reflect the systemic presence, self-reported data does not capture food and nutrient interactions ^38,39^ and the effects of food processing ^40^ that can affect systemic exposure.

When using nutritional biomarkers, it is important to consider interindividual differences in absorption, distribution, metabolism, and excretion (ADME) as potential limitations. In the case of the flavanol biomarkers used here, the impact of ADME has been previously addressed and discussed during the validation of gVLM_B_ and SREM_B_ as biomarkers ^26,27,31^. In this study, the impact of interindividual differences in ADME was also considered at the time of defining the thresholds levels of gVLM_B_ and SREM_B_, which was the basis for the classification of participants into the biomarker-based active and biomarker-based control groups. In this context, thresholds were defined as the lower 95% confidence level of gVLM_B_ and SREM_B_ level expected after the intake of 500 mg of CF (Figure 1), which aimed at reducing the risk of false negatives in the classification of participants into the biomarker-based active group. Sensitivity analyses confirmed the robustness of this approach as varying thresholds levels did not alter outcomes (Figure 4). Flavanol biomarker levels were assessed in spot urines and not 24 h urines, as spot urines are more commonly collected in large scale RCTs such as COSMOS. While the use of spot urines introduces uncertainty in flavanol biomarker level related to the time of sample collection, the combination of gVLM_B_ and SREM_B_ as complementary biomarkers helped addressing this limitation. In this context, gVLM_B_ and SREM_B_ have different systemic half-lives, for which the combination of these biomarkers can inform on both past and recent intakes. Nevertheless, gVLM_B_ and SREM_B_ better estimate intake around the day and time in which samples were collected. In this context, for a subset of participants we combined biomarkers levels estimated between 1- and 3-year follow-up, thus providing a wider time range for the biomarker-estimated flavanol intake assessment. Overall, nutritional biomarkers represent an objective and valid approach to assess flavanol intake, especially compared to self-report dietary and-pill intake questionnaires ^36,37^. Furthermore, biomarkers allow a better estimation of systemic exposure, which is key for the investigation of the health effect of dietary components. Similarly, the use of flavanol biomarkers facilitated the extrapolations of findings beyond the supplements tested in COSMOS. As shown in the case of biomarker-estimated assessment of the background diet, gVLM_B_ and SREM_B_ levels are also expected to increase after the consumption of flavanol-containing foods normally found in the diet, and thus, provide benchmarks for biomarkers levels that would be expected to mediate an effect.

We segmented participants into biomarker-based active and control groups, thus not relying on the original randomization of this study to create these two groups. This approach resulted in differences in baseline characteristics of participants in biomarker active and biomarker control groups at baseline (Table S2). However, adjusting for these baseline differences (model 3, Table S3) showed that this had little impact on estimated outcomes, and that biomarker-based segmentation still showed significant changes in HRs for all endpoints. Moreover, all were still larger than those determined via intervention-based ITT and PP analyses (Table 2).

COSMOS was used as a case study to explore the impact of background diet and adherence on RCTN outcomes. The findings from this work further confirm and strengthen the results reported in COSMOS main trial. Indeed, they suggest that there could be even stronger effects linked with flavanol intake than in the ITT and PP analysis in the COSMOS main trial ^25^. In addition, biomarker-estimated flavanol intake as part of background diet demonstrated that the amount of flavanols tested in COSMOS and similar to current recommendations ^24^ is achievable via a regular diet. In this context, further work will be needed to understand current flavanol intake levels in a US representative population. Such work will help assess the impact that increasing flavanol intake may have in the overall population.

## Conclusions

RCTN have an important role in the development of public health guidance, as they are considered the *gold standard* of evidence. In this study we have shown that the use of validated nutritional biomarkers is crucial to further strengthen the impact of RCTNs and increase scientific rigor and evidence quality. Including biomarker-based assessments in RCTNs requires the need of rethinking current study designs commonly implemented in these studies, including i) the trade-off between number of volunteers recruited and possibility of collecting samples in the totality of the cohort for biomarker analysis, ii) the possibility of extending ITT and PP analyses to include biomarker-based endpoints as pre-registered outcomes in RCTNs, and iii) consider biomarker levels at baseline as part of inclusion/exclusion criteria ^41^. At the same time, more validated nutritional biomarkers are critically needed both for individual dietary compounds and dietary patterns for integration into RCTN designs. This calls for further work aiming at the development and validation of nutritional biomarkers, which also includes investigating the ADME of dietary compounds under investigation and the analytical tools necessary to carry forward this research. There is a high cost in terms of funding, time and personnel needed to carry out RCTNs at scale. Their success in testing hypotheses relies strongly on knowledge of background diet and accuracy in assessing adherence to the intervention. More funding to develop and validate nutritional biomarkers would enhance the scientific significance and outcomes validity of RCTNs and subsequently their influence on diet and health recommendations and policy.

## Acknowledgments

We are deeply indebted to the 21,442 COSMOS participants for their steadfast and conscientious collaboration, and to the COSMOS Research Group for their scientific (Brigham and Women’s Hospital (BWH), Fred Hutchinson Cancer Research Center (FHCRC), Women’s Health Initiative (WHI), Data Safety and Monitoring Board (DSMB), Mars Edge) and logistical (BWH, FHCRC, DSMB, Mars Edge, Contract Pharmacal Corp, Pfizer Consumer Healthcare (now Haleon)) contributions.

## Statements and declarations

## Funding

COSMOS is supported by an investigator-initiated research grant from Mars Edge (JEM, HDS), a segment of Mars dedicated to nutrition research and products, which included infrastructure support and the donation of study pills and packaging. Pfizer Consumer Healthcare (now Haleon) provided support through the partial provision of study pills and packaging (JEM, HDS). COSMOS is also supported in part by grants AG050657, AG071611, EY025623, and HL157665 from the US National Institutes of Health (NIH), Bethesda, MD. The Women’s Health Initiative (WHI) program is funded by the National Heart, Lung, and Blood Institute, NIH, US Department of Health and Human Services through contracts 75N92021D00001, 75N92021D00002, 75N92021D00003, 75N92021D00004, and 75N92021D00005.

## Competing Interest Statement

J.I.O. and H.S. are employed by Mars, Incorporated, a company engaged in flavanol research and flavanol-related commercial activities. H.D.S. has received investigator-initiated research support from Mars Edge, Pure Encapsulations, and Pfizer Inc., and honoraria and/or travel for lectures from the Council for Responsible Nutrition, BASF, NIH, and American Society of Nutrition during the conduct of the study. J.E.M. has received investigator-initiated research support from Mars Edge. G.G.C.K. has received an unrestricted grant from Mars. COSMOS is supported by an investigator-initiated grant from Mars Edge, a segment of Mars dedicated to nutrition research and products, which included infrastructure support and the donation of study pills and packaging.

## Author Contributions

Designed research: JIO, GGCK, HS; performed research: JIO, GGCK; analyzed data: JIO, GGCK; wrote the paper: JIO, GGCK, HS, JWE, DMB, HDS, JEM

## Ethics approval

The COSMOS trial and ancillary studies were reviewed and approved by Institutional Review Boards at Brigham and Women’s Hospital/Mass General Brigham. The COSMOS website is www.cosmostrial.org. All participants provided informed consent. No identifiable information were used in this study.

## Notes

### Clinical Trial

NCT02422745

### Summary of Updates

Expanded on methodology and revised figures

