## Supplementary material for "The overlooked impact of background diet and adherence in nutrition trials"

^1^) Mars, Inc., McLean, VA; ^2^) Department of Pediatrics, Baylor College of Medicine, Houston, TX, USA ^3^) Department of Food Science and Human Nutrition, University of Illinois at Urbana-Champaign, Urbana-Champaign, IL, USA.^4^) Division of Preventive Medicine, Brigham and Women’s Hospital, Harvard Medical School. ^5^) Department of Food and Nutritional Sciences, University of Reading, Reading, UK;

Corresponding author: Gunter G. C. Kuhnle

### Supporting Information Text

#### Supplemental materials and methods

##### Study design

This study consisted of a post-hoc, secondary analysis of data emanated from a subcohort (n=6,532) of the COcoa Supplement and Multivitamin Outcomes Study (COSMOS, NCT 02422745), which is a recently completed randomized clinical trial in nutrition (RCTN) in 21,442 participants (including 8776 males ≥60 y and 12,666 females ≥65 y) in the US (1, 2). COSMOS followed a 2×2 factorial, randomized, double-blind, placebo-controlled design, and aimed at assessing the effects of cocoa extract (mainly containing cocoa flavanols) and a multivitamin supplement on total cardiovascular disease and total invasive cancer, respectively. The interventions included capsules containing cocoa extract that provided 500 mg/d cocoa flavanol (including 80 mg (–)-epicatechin) or a placebo (provided by Mars Edge) and a Centrum Silver daily multivitamin or placebo (provided by Pfizer Consumer Healthcare, now Haleon). For this study, we report the effects of the cocoa extract intervention.

The trial included women aged ≥65 y and men aged ≥60 y. Additional inclusion criteria were willingness to participate in the trial as evidenced by completion of informed consent and baseline forms. Major exclusion criteria were history of myocardial infarction or stroke; diagnosis of invasive cancer other than nonmelanoma skin cancer in the last 2 y prior to enrollment; serious illness that would preclude participation; taking cocoa extract, multivitamins, high-dose vitamin D, or high-dose calcium supplements and not willing to forego use during the trial; extreme sensitivity to caffeine; less than 75% compliance to study procedures during at least a 2-mo run-in period; and inability to communicate in English. COSMOS recruited participants from three main sources, including the Women’s Health Imitative (WHI), respondents to recruitment efforts for the VITamin D and OmegA-3 TriaL (NCT01169259) who were not ultimately randomized into the trial, and those identified through mass mailings and media efforts.

The population investigated in this study included a subcohort of 6,532 participants who provided spot urine samples during their participation in COSMOS [hereafter referred as COSMOS Biomarker cohort] (**Figure S1**). Urine samples were provided at baseline during the run-in phase of the study and at 1, 2 and/or 3-year follow-up. During the trial, participants completed a self-reported adherence assessment every 6 month by answering a series of questions related to the number of days taking study pills.

Enrollment period for the entire group of participants in COSMOS extended from April 2016 to March 2018, and the intervention was completed on December 31, 2020, with a median treatment period of 3.6 y. Further details of the protocol and main findings of the study were previously published [ref]. All participants provided written informed consent, and study approvals were obtained by the Institutional Review Board (IRB) at Mass General Brigham.

##### Flavanol biomarker quantification

Two flavanol biomarkers were assessed, including 5-(3ʹ,4ʹ-dihydroxyphenyl)-γ-valerolactone metabolites (gVLM_B_) and structurally related (–)-epicatechin metabolites (SREM_B_). gVLM_B_ represents the sum of two specific phase II metabolites of gut microbiota-derived flavanol catabolites, including 5-(4′-hydroxyphenyl)-γ-valerolactone-3′-sulfate and 5-(4′-hydroxyphenyl)-γ-valerolactone-3′-glucuronide. SREM_B_ represents the sum of three phase II flavanol metabolites, including (–)-epicatechin-3′-glucuronide, (–)-epicatechin-3′-sulfate and 3′-methyl-(–)-epicatechin-5-sulfate. Both, gVLM_B_ and SREM_B_ were previously validated as nutritional biomarkers and proved to provide complementary information about flavanol intake (3, 4). While gVLM_B_ informs on the intake of flavanols in general, SREM is a specific biomarker of the intake of (–)-epicatechin, one of the main bioactive flavanol present within the 500 mg/d CF tested in COSMOS (5). In addition, with gVLM_B_ having a longer systemic half-life than SREM (6), the combination of both biomarkers allows capturing different periods after flavanol intake and thus, increase the chance of identifying flavanol consumption in volunteers.

gVLM_B_ and SREM_B_ were quantified using validated methods based on UPLC-MS with authentic standards and corresponding ^13^C/^2^H-labeled internal standards (3, 4). Quality assurance was performed as described previously (7): each batch included two replicates of three performance quality control (QC) and one monitoring QC sample with known concentrations of biomarker. The acceptance criteria for each batch were that at least one performance QC at each concentration and four out of the six performance QCs were within 15% of the theoretical concentration. Concentrations outside the limit of quantifications were imputed by the values reported. This provides a better estimate of actual concentration than imputation by a fixed valued.

Flavanol biomarkers were measured in urine samples collected at baseline (n=6509), year 1 (n=1830), year 2 (n=1918) and year 3 (n=1050). Those volunteers who collected more than one urine sample at follow-up, corresponding concentrations of gVLM_B_ and SREM_B_ were averaged to simplify to one follow-up flavanol biomarker concentration per volunteer (n=2,051).

##### Adjustment of spot urine samples

Spot urine samples are often adjusted using urinary creatinine, although this method is likely to introduce bias (8). Specific gravity has been suggested as an alternative (9) and has been used previously (7), although it is not clear whether such an adjustment is always required. In this study we have therefore investigated a subset of urine samples (n=490) and determined specific gravity (1.02 (95% CI 1.01; 1.02); median (IQR)). Adjusting gVLM_B_ and SREM_B_ concentrations by specific gravity resulted in a median (IQR) concentration change of 0.05 (95% CI 0.01; 0.18) µmol/L or relative change of 1.5 (1.1; 2.0)%, but did not significantly alter ranking of participants (see Bland-Altman plot (10) in **Figure S2**). Thus, we determined that adjusting flavanol biomarkers levels not necessary for this study.

### Supplemental Figures

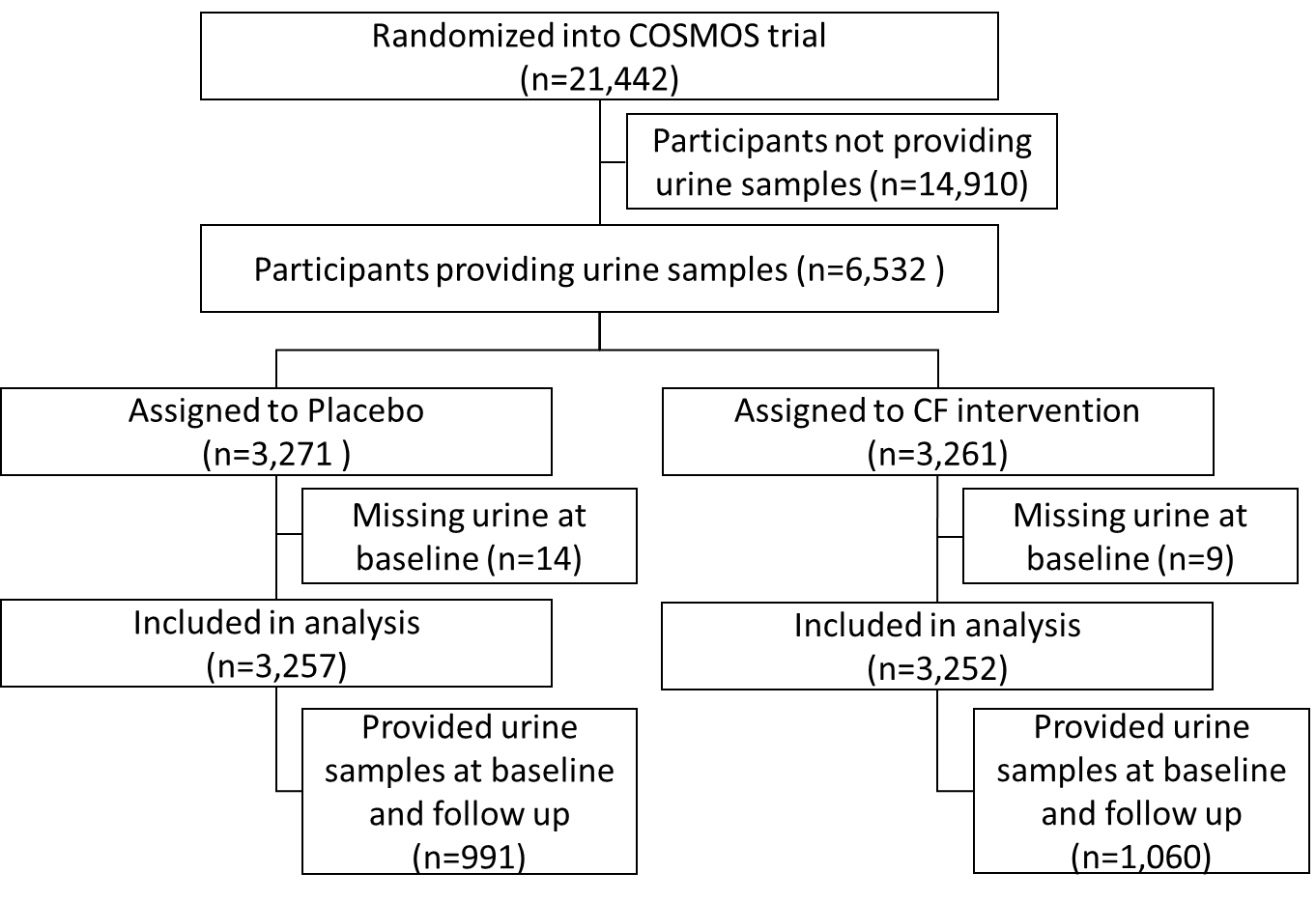

**Figure S1**. CONSORT Diagram of the COSMOS Biomarker Cohort

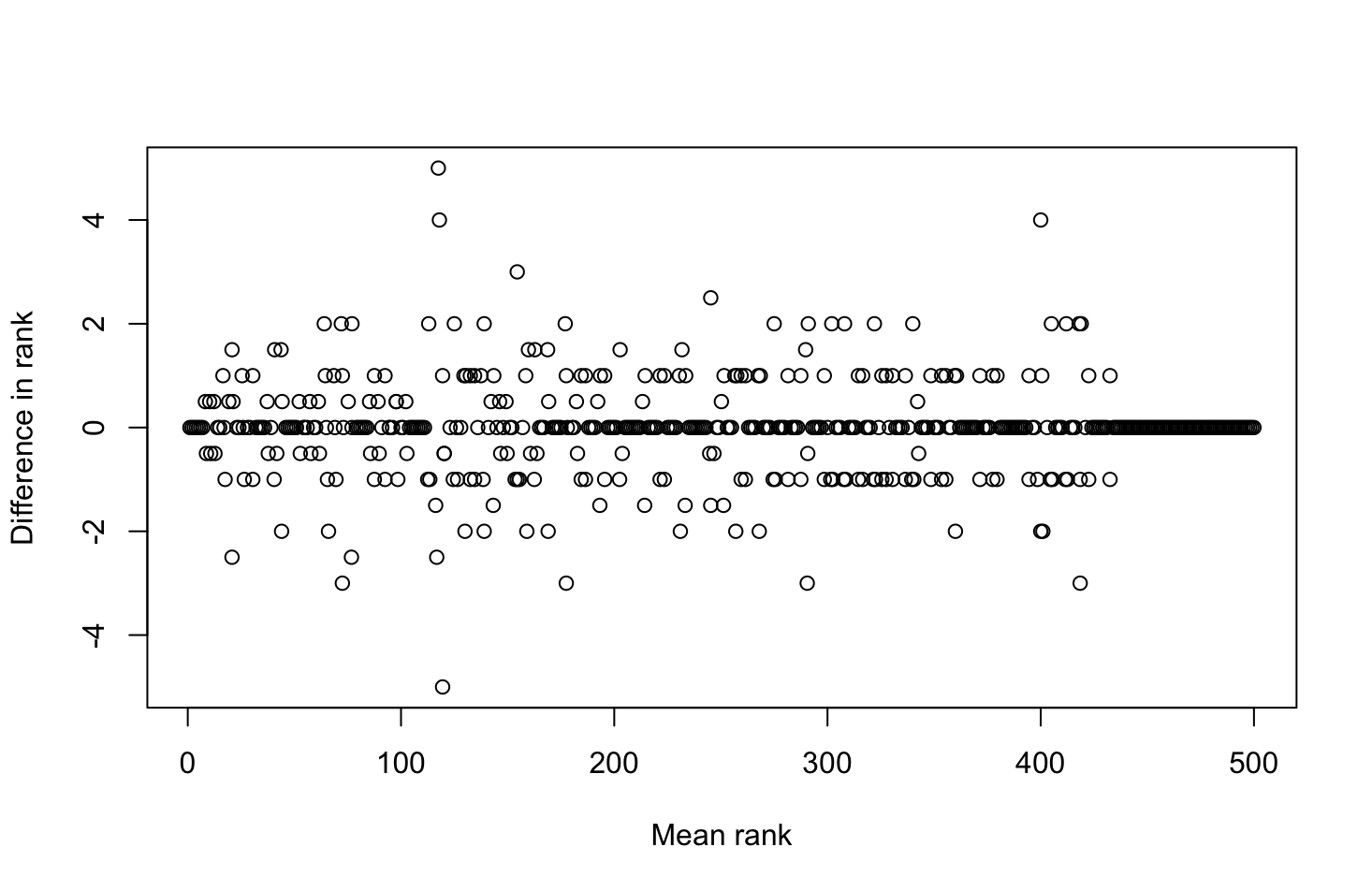

**Figure S2:** Bland-Altman plot (10) comparing participant rank of biomarker with and without adjustment for specific gravity. Difference in ranking of participants by flavanol biomarker concentrations in urine before and after adjusting by specific gravity.

### Supplemental Tables

**Table S1.** Characteristics of the study population. Data are mean ± SD or proportion. Test for difference between participants with and without urine samples using t-test or χ^2^ test.

|  | Total COSMOS Cohort | Participant who did not provide urine samples | Participant who provided urine samples  [COSMOS-Biomarker cohort] | | p |
| --- | --- | --- | --- | --- | --- |
| n | 21,442 | 14,910 | 6,532 | |  |
| Cocoa-flavanol intervention | 10,719 (50%) | 7458 (50%) | 3261 (50%) | | 0.908 |
| n of women | 12666 (59%) | 9325 (63%) | 3341 (51%) | | <0.001 |
| Age (y) | 72.1 ± 6.6 | 72.6 ± 6.6 | 71.0 ± 6.3 | | <0.001 |
| BMI (kg/m2) | 27.7 ± 5.4 | 27.7 ± 5.4 | 27.6 ± 5.3 | | 0.094 |
| Smoking status |  |  |  | | 0.317 |
| Never (n) | 11,565 (55%) | 8066 (55%) | 3499 (54%) | |  |
| Ever (n) | 8731 (41%) | 6018 (41%) | 2713 (42%) | |  |
| Current (n) | 835 (4%) | 591 (4 %) | 244 (4%) | |  |
| Participants from WHI cohort (n) | 4611 (22%) | 3639 (24%) | 972 (15%) | | <0.001 |
| Aspirin use (n) | 10379 (49%) | 7182 (49%) | 3197 (49%) | | 0.388 |
| Cholesterol medication use (n) | 9405 (44%) | 6480 (44%) | 2925 (45%) | | 0.115 |
| History of CVD (n) | 1269 (6%) | 882 (5.9) | 387 (5.9) | | 1.000 |
| History of hypertension (n) | 12423 (58%) | 8752 (59%) | 3671 (56%) | | 0.001 |
| CVD Risk factors (n) |  |  |  | | 0.914 |
| 0-1 | 9159 (43%) | 6376 (43%) | 2783 (43%) | |  |
| 2 | 6289 (29%) | 4358 (29%) | 1931 (30%) | |  |
| ≧3 | 5901 (28%) | 4101 (28%) | 1800 (28%) | |  |
| Diet quality (aHEI^†^, %) | 42.4 ± 11.0 | 42.3 ± 11.0 | 42.6 ± 10.8 | | 0.182 |
| **CVD Events** |  |  |  | |  |
| Total CVD events (n) | 866 (4%) | 609 (4%) | 257 (4%) | | 0.634 |
| CVD mortality (n) | 180 (1%) | 131 (1%) | 49 (1%) | | 0.386 |
| All-cause mortality (n) | 750 (3.5) | 556 (4%) | 194 (3%) | | 0.006 |
| Major cardiovascular events (n) | 559 (2.6) | 396 (3%) | 163 (3%) | | 0.527 |
| **Biomarker concentration (median [IQR])** | |  | |  |  |
| gVLM at baseline (µmol/L) | — | — | 3.3 [0.8, 11.0] | |  |
| gVLM at follow-up (µmol/L) | — | — | 8.0 [2.3, 23.2] | |  |
| SREM at baseline (µmol/L) | — | — | 0.6 [0.3, 2.0] | |  |
| SREM at follow-up (µmol/L) | — | — | 2.9 [0.8, 8.1] | |  |
| ^†^alternative Healthy Eating Index | | | | | |

**Table S2.** Participant characteristics of treatment groups and biomarker-based active and control groups. Data shown are mean ± SD or count and proportion. P-value is for test for difference between randomised groups and between biomarker-based groups using t-test or χ^2^ test.

|  | Randomised/Treatment assigned | |  | Biomarker-based | |  |
| --- | --- | --- | --- | --- | --- | --- |
|  | Placebo | Cocoa flavanols | p | Biomarker control | Biomarker active | p |
| n | 3271 | 3261 |  | 2,841 | 1,894 |  |
| n of participants randomized to CF pills (%) | 0 (0.0) | 3261 (100%) | **<0.001** | 298 (11%) | 1180 (62%) | **<0.001** |
| n of women (%) | 1679 (51%) | 1662 (51%) | 0.788 | 1529 (54%) | 841 (44%) | **<0.001** |
| Age (y) | 71.0± 6.3 | 71.0 ± 6.3 | 0.94 | 71.3 ± 6.3 | 70.3 ± 6.2 | **<0.001** |
| BMI (kg/m2) | 27.7 ± 5.3 | 27.5± 5.3 | 0.129 | 27.7 ± 5.4 | 27.4 ± 5.0 | **0.02** |
| Smoking status |  |  | 0.486 |  |  | 0.516 |
| Never (n) | 1741 (54%) | 1758 (55%) |  | 1519 (54%) | 1022 (55%) |  |
| Ever (n) | 1363 (42%) | 1350 (42%) |  | 1176 (42%) | 788 (42%) |  |
| Current (n) | 131 (4%) | 113 (4%) |  | 116 (4%) | 65 (4%) |  |
| Participants from WHI cohort | 504 (15%) | 468 (14%) | 0.244 | 454 (16%) | 220 (12%) | **<0.001** |
| Aspirin use (n) | 1592 (49%) | 1605 (50%) | 0.701 | 1384 (49%) | 942 (50%) | 0.579 |
| Cholesterol medication use (n) | 1448 (45%) | 1477 (45%) | 0.528 | 1271 (45%) | 828 (44%) | 0.413 |
| History of CVD (n) | 207 (6%) | 180 (6%) | 0.183 | 184 (7%) | 100 (5%) | 0.102 |
| History of hypertension (n) | 1850 (57%) | 1821 (56%) | 0.529 | 1637 (58%) | 1016 (54%) | **0.006** |
| CVD risks |  |  | 0.227 |  |  | **0.004** |
| 0-1 (n) | 1382 (42%) | 1401 (43%) |  | **1181 (47%)** | **852 (45%)** |  |
| 2 (n) | 947 (29%) | 984 (30%) |  | **820 (29%)** | **570 (30%)** |  |
| ≧3 (n) | 931 (29%) | 869 (27%) |  | **828 (29%)** | **472 (25%)** |  |
| Diet quality (aHEI†, %) | 42.7 ± 11.0 | 42.5 ± 10.7 | 0.538 | 42.4 ± 10.9 | 43.3 ± 10.8 | **0.007** |
| CVD events |  |  |  |  |  |  |
| Total CVD events (n) | 141 (4%) | 116 (4%) | 0.133 | 128 (5%) | 56 (3%) | **0.009** |
| CVD mortality (n) | 32 (1%) | 17 (0.5%) | **0.046** | 29 (1%) | 8 (0%) | **0.034** |
| All-cause mortality (n) | 108 (3%) | 86 (3%) | 0.131 | 100 (4%) | 34 (2%) | **0.001** |
| Major CVD events (n) | 94 (3%) | 69 (2%) | 0.050 | 89 (3%) | 28 (2%) | **<0.001** |
| Biomarker concentration (median [IQR]) | | | | | | |
| gVLM baseline (µmol/L) | 3.4 [0.8, 11.1] | 3.21 [0.8, 10.9] | 0.49 | 2.0 [0.5, 5.7] | 20.9 [4.6, 36.2] | **<0.001** |
| gVLM follow-up (µmol/L) | 4.2 [1.4, 10.9] | 16.04 [5.1, 34.9] | **<0.001** | 3.2 [1.0, 7.3] | 22.5 [8.5, 37.7] | **<0.001** |
| SREM baseline (µmol/L) | 0.6[0.3, 1.8] | 0.61 [0.3, 2.1] | 0.187 | 0.4 [0.3, 1.2] | 1.5 [0.4, 5.3] | **<0.001** |
| SREM follow-up (µmol/L) | 1.0 [0.4, 2.4] | 7.41 [3.3, 11.5] | **<0.001** | 1.1 [0.4, 2.8] | 7.9 [2.6, 11.6] | **<0.001** |
| †alternative Healthy Eating Index | | | | | | |
